# FASDetect – A machine learning-based app to screen for the risk of fetal alcohol-spectrum disorder in youth with attention-deficit/hyperactivity disorder symptoms

**DOI:** 10.1101/2022.09.12.22279880

**Authors:** Lukas Ehrig, Ann-Christin Wagner, Heike Wolter, Christoph U. Correll, Olga Geisel, Stefan Konigorski

**Affiliations:** Digital Health Center, Hasso Plattner Institute for Digital Engineering, University of Potsdam, Potsdam, Germany; Department of Child and Adolescent Psychiatry, Charité Universitätsmedizin Berlin, Berlin, Germany; The Zucker Hillside Hospital, Department of Psychiatry, Northwell Health, Glen Oaks, NY, USA; Donald and Barbara Zucker School of Medicine at Hofstra/Northwell, Department of Psychiatry and Molecular Medicine, Hempstead, NY, USA; Hasso Plattner Institute for Digital Health at Mount Sinai, Icahn School of Medicine at Mount Sinai, New York, NY, USA; Department of Statistics, Harvard University, Cambridge, MA, USA

**Author notes:** Correspondence: Stefan Konigorski, PhD, Digital Health Center, Hasso Plattner Institute for Digital Engineering, Rudolf-Breitscheid-Str. 187, 14482 Potsdam, Germany. these authors contributed equally to the article (Lukas Ehrig, Ann-Christin Wagner). these authors contributed equally to the article (Olga Geisel, Stefan Konigorski).

## Abstract

Fetal alcohol-spectrum disorder (FASD) is underdiagnosed and often misdiagnosed as attention-deficit/hyperactivity disorder (ADHD). Here, we developed a screening tool for FASD in youth with ADHD symptoms. To develop the prediction model, medical record data from a German University outpatient unit were assessed including 275 patients aged 0-19 years old with FASD with or without ADHD and 170 patients with ADHD without FASD aged 0-19 years old. We trained 6 machine learning models based on 13 selected variables and evaluated their performance. Random forests yielded the best prediction models with a cross-validated AUC of 0.92 (95% confidence interval [0.84, 0.99]). Follow-up analyses indicated that a random forest model with 6 variables – body length and head circumference at birth, IQ, socially intrusive behaviour, poor memory and sleep disturbance – yielded equivalent predictive accuracy. We implemented the prediction model in a web-based app called FASDetect – a user-friendly, clinically scalable FASD risk calculator that is freely available at https://fasdetect.dhc-lab.hpi.de.

## Introduction

Fetal alcohol-spectrum disorder (FASD) is an umbrella term for medical conditions caused by prenatal alcohol exposure, including fetal alcohol syndrome (FAS), partial fetal alcohol syndrome (pFAS), alcohol related birth defects (ARBD), and alcohol-related neurodevelopmental disorder (ARND). The global prevalence of FASD is estimated to be between 2-5% of the Western world’s population (1). Despite the prevalence rate, FASD is highly underdiagnosed and many patients miss out on the beneficial effects of an early childhood diagnosis and subsequent early intervention (2-5).

Established diagnostic systems for FASD are based on the manifestation of growth deficiencies, craniofacial dysmorphia, central nervous system damage/dysfunction, and gestational alcohol exposure (6, 7). These neuropsychological impairments can manifest as deficits in intelligence, learning, memory, executive function and academic achievements, language and motor development and attention (8). People with FASD have a higher risk to develop secondary psychiatric conditions, like conduct disorder, attention-deficit/hyperactivity disorder (ADHD) and sleep disorders, as well as to experience adverse life events (8-11). Hyperactivity, inattention and impulsivity are characteristically seen both in patients with ADHD and FASD. More than half of FASD patients suffer from comorbid ADHD (11). These overlapping symptoms of FASD and ADHD complicate the diagnostic process and can lead to misdiagnosis as well as delayed intervention for FASD. In a study conducted in 547 children and adolescent who were adopted or in foster care and who underwent a comprehensive multidisciplinary diagnostic evaluation to identify FASD, 156 youth met criteria for FASD, but as many as 87% of them had been misdiagnosed, most commonly with ADHD, or had remained undiagnosed (12). The very high misdiagnosis rates underscore the importance of evaluating youth diagnosed with ADHD in order to detect any missed FASD diagnosis.

The purpose of the present study was to (i) develop a machine learning algorithm for detection of FASD in patients with ADHD symptoms based on retrospectively gathered out-patient data, and (ii) subsequently use this algorithm to create an easy and fast as well as clinically scalable online screening tool. We hypothesized that we would be able to identify an algorithm with sufficient accuracy to differentiate youth with versus without FASD.

## Results

### Study sample

This study was conducted at the outpatient unit of the department of child and adolescent psychiatry at the Campus Charité Virchow of the Charité Universitätsmedizin Berlin, Germany. For the analysis, a group of consecutively assessed patients with a clinical diagnosis of ADHD without FASD and a group of patients with an expert diagnosis of FASD (with or without comorbid ADHD) was compared. Altogether, 694 patients with ADHD symptoms were identified consecutively from the general patient pool being potentially eligible for the study. 256 of the 694 ADHD patients had a confirmed FASD diagnosis and therefore were excluded from the ADHD pool. Further, 141 patients were excluded from the ADHD group due to an unconfirmed ADHD diagnosis; 58 because they had a suspected but not confirmed FASD diagnosis; 37 due to other severe medical, psychiatric, or neurological conditions; and 32 patients were excluded because patient records were unavailable. This yielded in total 170 patients in the ADHD group. The consecutively enrolled FASD group was recruited from the specialist center and consisted of 275 youth, including 129 FASD patients with comorbid ADHD and 146 patients without comorbid ADHD diagnosis. These 275 patients included most of the 256 FASD patients from the general patient pool.

### Description of the patients’ characteristics

Tables 1-2 give an overview of the main characteristics of the n = 445 FASD and ADHD patients, which included 159 female (mean age at initial presentation, 9.6 years [range, 0.2–18.8 years]) and 286 male (mean age at initial presentation, 8.9 years [range, 0.1–19.0 years]) patients. 139 of the FASD patients had a FAS diagnosis, 127 had a pFAS diagnosis and 9 patients were diagnosed with ARND. 170 patients belonged to the ADHD group (31 female; mean age at initial presentation, 8.7 years [range, 3.7–16.8 years]; 139 male; mean age at initial presentation, 8.4 years [range, 2.3–15.7 years]) and 275 patients belonged to the FASD group (128 female; mean age at initial presentation, 9.9 years [range, 0.2–18.8 years]; 147 male; mean age at initial presentation, 9.4 years [range, 0.1–19.0 years]). There were very low pairwise correlations between these variables, with the exception of head circumference and birth length (Pearson correlation coefficient of 0.57).

**Table 1:** Mean and standard deviation (SD) of number of mothers pregnancies and births, gestational age, age at first presentation, length, weight, head circumference at time of birth and first presentation for all patients, FASD and ADHD patient groups.

| Variable | All patients |  | FASD patients |  | ADHD patients |  |
| --- | --- | --- | --- | --- | --- | --- |
|  | Mean | SD | Mean | SD | Mean | SD |
| age at first presentation | 9.15 | 4.03 | 9.60 | 4.53 | 8.43 | 2.94 |
| gestational age | 38.57 | 2.39 | 38.58 | 2.21 | 38.56 | 2.68 |
| z-score of birth length | -0.58 | 1.16 | -0.87 | 1.13 | -0.08 | 1.03 |
| z-score of birth weight | -0.48 | 1.19 | -0.80 | 1.09 | 0.05 | 1.17 |
| z-score of head circumference at birth | -0.45 | 1.11 | -0.70 | 1.11 | 0.07 | 0.93 |
| z-score of length at first presentation | -0.27 | 1.17 | -0.50 | 1.12 | 0.27 | 1.12 |
| z-score of weight at first presentation | -0.02 | 1.21 | -1.20 | 1.15 | 0.38 | 1.24 |
| z-score of head circumference at first presentation | -1.06 | 1.55 | -1.37 | 1.37 | 0.23 | 1.58 |
| number of mothers pregnancies | 2.78 | 2.14 | 2.84 | 2.12 | 2.53 | 2.22 |
| number of mothers births | 2.09 | 1.39 | 2.21 | 1.48 | 1.91 | 1.23 |

**Table 2:** Absolute and relative frequencies for sex and cognitive, behavioral and sleep variables, speech development, psychiatric comorbidities and medication in all patients, FASD and ADHD patient groups

| Variable | All patients |  | FASD patients |  | ADHD patients |  |
| --- | --- | --- | --- | --- | --- | --- |
|  | Yes | No | Yes | No | Yes | No |
| male sex | 286<br>(64.3%) | 159<br>(35.7%) | 147<br>(53.5%) | 128<br>(46.5%) | 139<br>(81.8%) | 31<br>(18.2%) |
| Intelligence below 85 IQ points | 119<br>(34.4%) | 227<br>(65.6%) | 92<br>(51.4%) | 87<br>(48.6%) | 27<br>(16.2%) | 140<br>(83.8%) |
| socially intrusive behaviour | 196<br>(47.6%) | 216<br>(52.4%) | 155<br>(63.5%) | 89<br>(36.5%) | 41<br>(24.4%) | 127<br>(75.6%) |
| Impairment in memory | 272<br>(66.2%) | 139<br>(33.8%) | 208<br>(86.0%) | 34<br>(14.0%) | 64<br>(37.6%) | 105<br>(62.1%) |
| sleep disorder/disturbance | 147<br>(37.1%) | 249<br>(62.9%) | 107<br>(47.3%) | 119<br>(52.7%) | 40<br>(23.5%) | 130<br>(76.5%) |
| speech development disorder | 221<br>(54.2%) | 187<br>(45.8%) | 145<br>(60.7%) | 94<br>(39.3%) | 76<br>(45.0%) | 93<br>(55.0%) |
| psychiatric comorbidities | 289<br>(72.3%) | 111<br>(27.8%) | 185<br>(80.4%) | 45<br>(19.6%) | 104<br>(61.2%) | 66<br>(38.9%) |
| prescribed psycho-tropic medications | 186<br>(47.7%) | 204<br>(52.3%) | 64<br>(29.1%) | 156<br>(70.9%) | 122<br>(71.8%) | 48<br>(28.2%) |

### Prediction models to separate FASD and ADHD

The statistical analysis aimed at developing and evaluating a prediction model that would be able to separate FASD from ADHD cases with sufficient accuracy. After data preprocessing and variable selection (see Materials & Methods), we tested the performance of 6 machine learning algorithms to predict ADHD or FASD using the 13 remaining variables on our data with nested cross-validation. Table 3 provides an overview of the main results for the prediction model based on the 13 variables number of mother’s births, gestational age, z-scores of length, weight and head circumference at birth, z-scores of length and weight at initial presentation, as well as the presence of low IQ, socially intrusive behavior, speech development disorder, poor memory, sleep disturbance and psychiatric comorbidities. When predicting FASD cases among ADHD patients, an AUC of 0.92 (95% confidence interval CI [0.84, 0.99]) was reached by the RF model. 91% of the FASD patients were correctly identified and overall 85% of patients received a correct classification. Of all patients that were classified as FASD cases, 86% were true FASD cases. The kNN and Gaussian Process classifiers both reached an AUC of 0.90 and accuracy of 0.84. The SVM also had a ROC AUC of 0.90 ([0.80, 0.99]), but recognized more positive cases with a sensitivity of 0.92, the highest among all evaluated algorithms. Logistic regression and GBDT both yielded an AUC of 0.91 (95% CI [0.83, 0.99] and 0.91 [0.82, 0.99], respectively). The highest positive predictive value (0.89) was reached by the logistic regression model, however at the cost of the lowest sensitivity (0.84). The RF had a Brier score of 0.11, the other models had a Brier score of 0.12.

**Table 3:** Cross-validated evaluation results of prediction with 13 variables. The best model is highlighted in italics.

| Model (13 variables) | AUC | Accuracy | Precision | Recall | Brier |
| --- | --- | --- | --- | --- | --- |
| Logistic Regression | 0.91 [0.83, 0.99] | 0.84 | 0.89 | 0.84 | 0.12 |
| Support Vector Machine | 0.90 [0.80, 0.99] | 0.85 | 0.85 | 0.92 | 0.12 |
| *Random Forest* | *0.92 [0.84, 0.99]* | *0.85* | *0.86* | *0.91* | *0.11* |
| Gradient Boosting Decision Tree | 0.91 [0.82, 0.99] | 0.85 | 0.86 | 0.91 | 0.12 |
| kNN Classifier | 0.90 [0.81, 0.99] | 0.84 | 0.87 | 0.88 | 0.12 |
| Gaussian Process Classifier | 0.90 [0.81, 0.99] | 0.84 | 0.86 | 0.89 | 0.12 |

In all experiments and cross-validation trials, only 6 of the 13 variables were frequently selected in the ML pipelines. These six variables were: z-scores of body length and head circumference at birth, IQ below 85 IQ points, socially intrusive behaviour, poor memory and sleep disturbance. When using this reduced variable set in our second set of analyses, the RF model had an AUC of 0.93 (95% CI [0.85, 1]) and could on average identify 91% of FASD cases in the test sets, with 85% of patients being classified correctly. Patients that were classified as FASD patients were true cases in 87%. All other algorithms separated the ADHD and the FASD groups similarly well with an AUC of 0.90 or 0.91 (see Table 4). Hence, the various performance metrics of the algorithms were very similar compared to the prediction models using 13 variables. The results of all experiments including ROC curves can be found in Supplementary Figures S1-S6.

**Table 4:** Cross-validated evaluation results of the pipeline with 6 variables. The best model is highlighted in italics.

| **Model (6 variables)** | **AUC** | **Accuracy** | **Precision** | **Recall** | **Brier** |
| --- | --- | --- | --- | --- | --- |
| Logistic Regression | 0.90 [0.81, 0.99] | 0.82 | 0.89 | 0.81 | 0.13 |
| Support Vector Machine | 0.90 [0.81, 0.99] | 0.84 | 0.85 | 0.91 | 0.12 |
| *Random Forest* | *0.93 [0.85, 1.00]* | *0.85* | *0.87* | *0.91* | *0.11* |
| Gradient Boosting Decision Tree | 0.91 [0.83, 0.99] | 0.84 | 0.87 | 0.87 | 0.12 |
| kNN Classifier | 0.91 [0.83, 0.99] | 0.84 | 0.87 | 0.88 | 0.11 |
| Gaussian Process Classifier | 0.91 [0.82, 0.99] | 0.83 | 0.86 | 0.88 | 0.12 |

For our screening application, we selected the RF model because of its high sensitivity, robustness to changes of the variable set, and its good overall performance. The probability score distributions of the RF model are depicted in Figure 2 and illustrate that the estimated probabilities of having FASD are generally high for FASD patients and low for ADHD patients. There are only few patients that are assigned a low risk of FASD while having a diagnosis of FASD or that are assigned a high risk despite having an ADHD diagnosis without FASD. The figure also shows the number of true and false classifications at different probability thresholds. For any probability threshold used for the decision whether a patient is assigned to the ADHD or FASD group, ADHD patients right of that threshold (i.e., that were assigned a higher probability by the prediction model) are false positives, FASD patients left of that threshold are false negatives, and all others were classified correctly.

### Implementation of machine learning model in FASDetect screening application

In the final step, we developed a screening app for the detection of FASD among ADHD cases based on the RF algorithm. Our focus was on the target user group of medical professionals from different fields (e.g., pediatricians, psychiatrists). Requirements derived for the application included that it should be user-friendly, quick and easy-to-use and that the screening result is immediately visible.

The frontend of the application was built using Vue.js/quasar, the backend using Python/flask. The resulting app consists of three screens and is based on the RF model of 6 variables that can be quickly and appropriately assessed by all possible users. The first screen contains the disclaimer and provides some information about the app. The next screen contains a questionnaire, where information about the 6 variables is obtained. The last screen shows the results and some context of how to interpret the screening results (see Figure 4). In order to facilitate quick decision making, the results are visually represented using a traffic light metaphor. A yellow signal is shown in FASDetect when the model estimates the FASD risk to be 50-74% and therefore classifies the patient as a potential FASD case. When the risk exceeds 75%, the red signal is shown, indicating a high risk. The FASDetect app is designed in such a way that if all the variables are known, the data entry and retrieval of the result can be completed very easily in less than 1 minute. Currently, the app exists in English and German, but can easily be extended to include more languages. The app is available open-source and free-of-charge at https://fasdetect.dhc-lab.hpi.de.

## Discussion

Diagnosis of FASD is often challenging as well as time-consuming and the most common mental health diagnosis given to FASD patients is ADHD when missing the FASD diagnosis (12). Currently, there exists no tool to screen the risk of FASD in ADHD patients. In our study, we created a tool called FASDetect to detect FASD among patients with ADHD symptoms. Health care providers need to elicit information to answer 6 questions to receive a quick screening result.

FASDetect is based on machine learning models. In our study, we compared different machine learning algorithms and implemented random forest in FASDetect, which performed best with an AUC of 0.92. This predictive accuracy is similar to previous studies using machine learning in ADHD. Duda et al. showed that machine learning algorithms are capable of accurately differentiating between patients with ADHD and autism-spectrum disorder with a similar AUC of 0.965 (18). Zhang et al. successfully used machine learning to distinguish between FASD patients and healthy controls through use of eye movement, psychometric and neuroimaging data with 85% classification accuracy (19). Our work emphasizes the great potential of machine learning to optimize screening and diagnostic procedures that can help improve treatment selection and outcome predictions in clinical psychology and psychiatry (20).

The 6 most important variables that were retained for efficient FASD screening via FASDetect are the z-scores of birth length and birth head circumference, low IQ, social intrusiveness, poor memory and sleep disturbance. All of these variables are known to be medically linked to FASD (21-27). Previous studies have shown that FASD patients are more likely to be microcephalic and remain to be microcephalic and length growth-restricted throughout life. They also show a lower intelligence than ADHD patients and have been found to suffer from memory problems. Socially intrusive behaviour and sleep disturbance are also often seen in FASD patients. All of this is also shown in our data, which adds face validity to the finding that these predictors were selected during automatic feature selection. Thus, we are optimistic that our results will generalize and can be replicated in other populations.

FASDetect may represent the time-saving clinical screening application for FASD that has been missing until now. Such a tool is urgently needed in clinical practice. In next steps, FASDetect has to be evaluated prospectively and licensed for any medical use. Then, we can imagine the following use: If the screening result shows red or yellow, further medical examination is highly recommended. Child psychiatrists who specialise in FASD should examine the patient and investigate the presence of FASD. Experts consider additional information, such as facial dysmorphia or prenatal alcohol exposure that are required to meet official medical diagnostic criteria but were considered inapplicable for a screening tool.

Paediatricians vastly underrecognize FASD and are often unfamiliar with the diagnostic criteria, leading to a higher chance for misdiagnosis and missed diagnosis (28). The risk of underrecognition and misdiagnosis is at least as high for child and adolescent psychiatrists. FASDetect could enable inexperienced medical staff to screen for FASD and direct patients to specialists. This can help FASD patients to be diagnosed earlier in life and be seen by specialists. Thus, FASDetect could help to reduce the misdiagnosis rates and aide the diagnostic process in busy clinical settings. The successful implementation promises an earlier diagnosis for FASD patients who are currently frequently incorrectly diagnosed with ADHD. Thus, patients who are screened using FASDetect will benefit from earlier treatment, a reduction of secondary conditions and eventually from improved general health.

### Limitations

The results of this study have to be interpreted within its limitations. First, the analysis of archived patient records was limited by the available content of the data. Including further clinical variables might further improve the predictive accuracy of FASDetect. Second, we only examined the discriminatory power and accuracy of the FASDetect app for FASD cases among a sample of patients with a primary diagnosis of ADHD. Further studies are needed that include a broader variety of mental health diagnoses, ideally also oppositional defiant disorder, autism-spectrum disorders and youth with intellectual disability/low IQ who share some other features of FASD than patients with ADHD. The inclusion of further variables that were not available such as reduced eyesight, head circumference at initial presentation and academic achievements are promising predictor candidates for future iterations of the model that are relatively easy to obtain clinically and that should therefore be assessed in future studies.

Third, FASD cases were not distributed evenly within the spectrum (139 FAS, 127 pFAS, 9 ARND), which may have aided the differentiation of the ADHD and FASD groups by the machine learning algorithms. Future research is needed to evaluate how well FASDetect identifies patients across the entire FASD spectrum. Fourth, the study was conducted in a university setting, and testing of generalizability to other clinical settings is further required. Fifth, the patient data for the FASD cases was gathered by psychiatrists specialized in FASD diagnosis. The ADHD cases were diagnosed by outpatient clinicians trained in child and adolescent psychiatry, but without a specific focus on ADHD. The high level of expertise and elaborate testing (e.g. intelligence testing) cannot necessarily be expected of the average user of FASDetect. We adapted the selection of variables that went into final screening tool accordingly. Nevertheless, it is possible that variables seem less distinctive to lesser experienced pediatricians and may be underrecognized when screening with FASDetect.

To our knowledge, this study is the first that developed an empirically-based, machine-learning-derived screening app that robustly differentiates between FASD and ADHD using parameters that can be relatively easily obtained as part of clinical care. The tool, which we call FASDetect, provides a green-yellow-red light rating system on the risk for FASD in ADHD patients calculated from easily obtainable patient data and is an efficient tool for general pediatric practice. The FASDetect is freely available, and we hope that future research with this tool can validate and extend its utility and assess to what degree FASDetect can aide clinical diagnosis and decision making for subjects with FASD compared to usual care.

## Materials and Methods

### Study population

This study was conducted at the outpatient unit of the department of child and adolescent psychiatry at the Campus Charité Virchow of the Charité Universitätsmedizin Berlin, Germany. For the analysis, a group of consecutively assessed patients with a clinical diagnosis of ADHD without FASD and a group of patients with an expert diagnosis of FASD (with or without comorbid ADHD) was compared. ADHD patients were included from the general pool of patients who were treated at Campus Charité Virchow of the Charité Universitätsmedizin Berlin between January 2019 and September 2020. FASD patients were included from two sources: from the general pool of ADHD patients described above, as well as from the pool of ambulatory patients of the FASD specialist center at the Campus Charité Virchow of the Charité – Universitätsmedizin Berlin who were treated between January 2019 and September 2020. The two groups were ascertained based on the following inclusion and exclusion criteria.

Inclusion criteria for children and adolescents with ADHD were

a. age between 0 and 19 years,
b. diagnosis of ADHD, combined type of inattentive type, with or without oppositional defiant or conduct disorder according to ICD-10 by child and adolescent psychiatrists at our department of child and adolescent psychiatry at the Campus Charité Virchow of the Charité Universitätsmedizin Berlin,
c. diagnosis of ADHD confirmed during longitudinal assessment and care at our department

Exclusion criteria for children and adolescents with ADHD were

a. severe medical, psychiatric, or neurological conditions (such as microdeletion, microduplication, genetic syndromal diseases, autism-spectrum disorders or hydrocephalus) which can affect the youth’s behaviour
b. suspected or confirmed comorbid FASD diagnosis

Inclusion criteria for children and adolescents with FASD (with or without ADHD) were

a. age between 0 and 19 years,
b. diagnosis of FASD according to ICD-10 and the 4 digit code (7)
c. diagnosis of FASD confirmed as part of longitudinal assessment and care at our department

Exclusion criteria for children and adolescents with FASD were severe medical, psychiatric, or neurological conditions.

Of each patient, the following data were extracted retrospectively from medical records: height, weight and head circumference at all available time points; presence or absence of any psychiatric comorbidities, prescribed psychotropic medications yes versus no, fascial dysmorphia and malformation; the results of intelligence tests, whether or not the patient’s IQ was below 85 IQ points; as well as pregnancy- and birth-related data such as consumption of alcohol, nicotine and other drugs, number of the mother’s pregnancies and births, child’s gestational age at first ultrasound and at time of birth, Apgar score (13) and pH of the umbilical cord after birth. The presence or absence of oppositional, hyperactive and impulsive behavior, lack of concentration and attention, developmental disorders, sleep disorders, socially intrusive behavior, and impaired executive function and cognitive flexibility were also assessed clinically. Those symptoms were recorded during clinical assessments, history taking, parent and patient interviews and through behavioral questionnaires such as the child behavior checklist (14) or DISYPS (15). Assessed symptoms were documented as “present” or “absent”, no degree of severity was assessed.

### Statistical analysis

The statistical analysis aimed at developing and evaluating a prediction model that would be able to separate FASD from ADHD cases with sufficient accuracy. All machine learning analyses were performed in Python 3.7.3. The code is publicly available at https://github.com/HIAlab/FASDetect. After overall data quality control steps, the training and evaluation of different prediction algorithms was performed in several steps.

In a first overall quality control step, we removed variables with more than 35% missing values for either group (ADHD/FASD). This missing values threshold was chosen in order to include head circumference at birth, which had 35% missing values for ADHD patients, as an indicator for growth deficiencies in FASD patients that is easy to assess and well-suited for use in a screening application. The quality control retained 42 predictive variables, from which we further removed variables with redundant information, such as re-coded duplicates (20 variables), variables that would be too complex to assess for practitioners during a clinical screening visit (5 variables, e.g., executive dysfunction), and variables that might limit generalizability (8 variables). For some variables, multiple reasons for exclusion applied. From the resulting 13 variables, none had more than 23% missing values across both the ADHD and FASD groups. On average, 11% of the variable values were missing for the ADHD group and 12% for the FASD group.

Next, we tested the performance of 6 machine learning algorithms to predict ADHD or FASD using the 13 remaining variables on our data with nested cross-validation (see Figure 1). To initialize our machine learning pipeline, we randomly split the entire data set into 10 folds (outer split), where each of these folds consisted of 10% of the ADHD cases (n = 170) and 10% of the FASD cases (n = 275), respectively. We used these outer folds to perform 10-fold cross-validation (CV) with nine folds for training and the remaining fold for testing. The training data from the outer split with 90% of the data were split again into 10 stratified folds used for training and 10% for validation of the hyperparameters of the pipeline (see below) using a grid search. After the optimized hyperparameter configuration was found in the nested 10-fold CV, the respective model was refit on the complete training data of the outer split (i.e. training and validation data of the inner split) and evaluated against the fold’s test set. The nested CV scheme is depicted in Figure 1.

**Figure 1:**
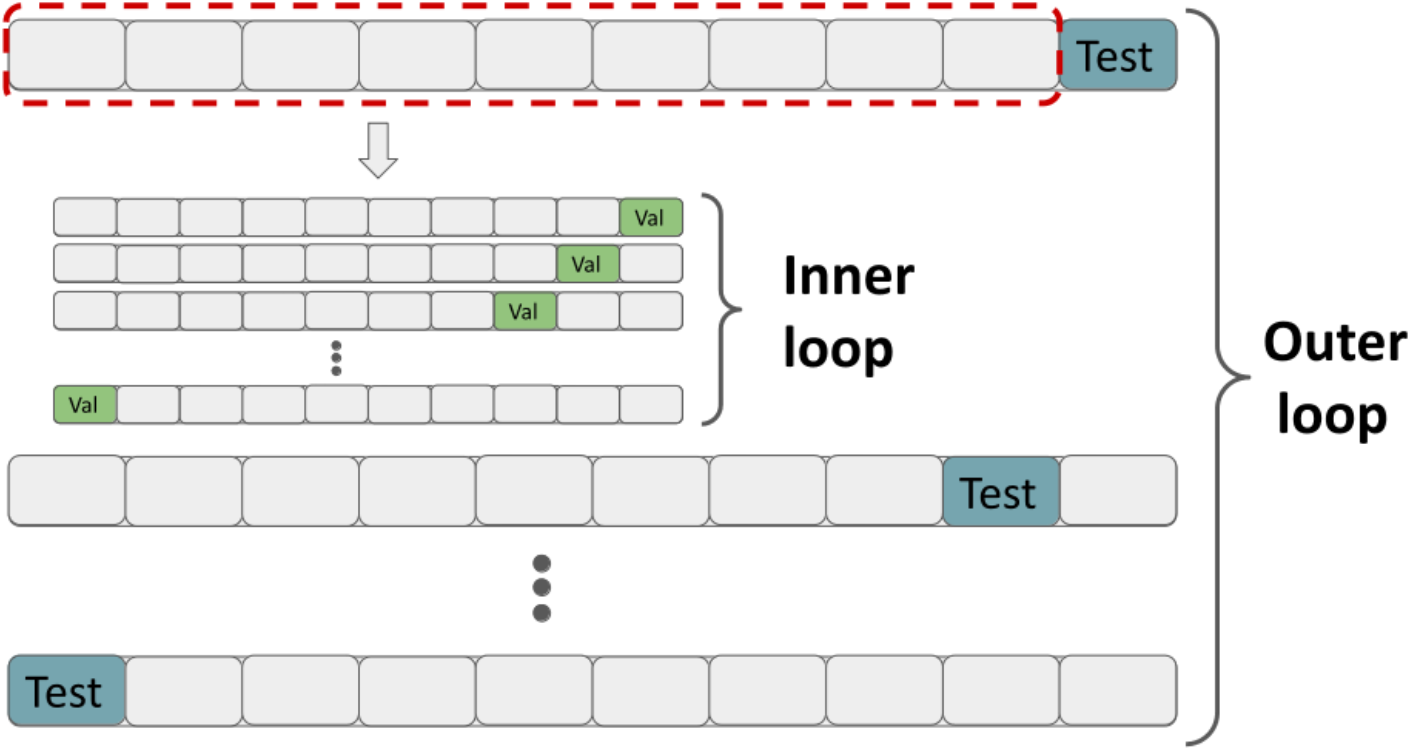
Overview of the 10-fold nested cross-validation procedure. The data are randomly split into 10 stratified folds where one fold is held out as a test set (blue). For each split, the 9 folds are split again into 10 folds, with one fold (green) to validate the hyperparameters. The hyperparameters with the best average ROC AUC on the validation sets are used to fit the machine learning pipeline on the complete training set (i.e., the 9 outer folds framed in red) and tested against the test set (blue), resulting in 10 ROC AUC scores.

**Figure 2:**
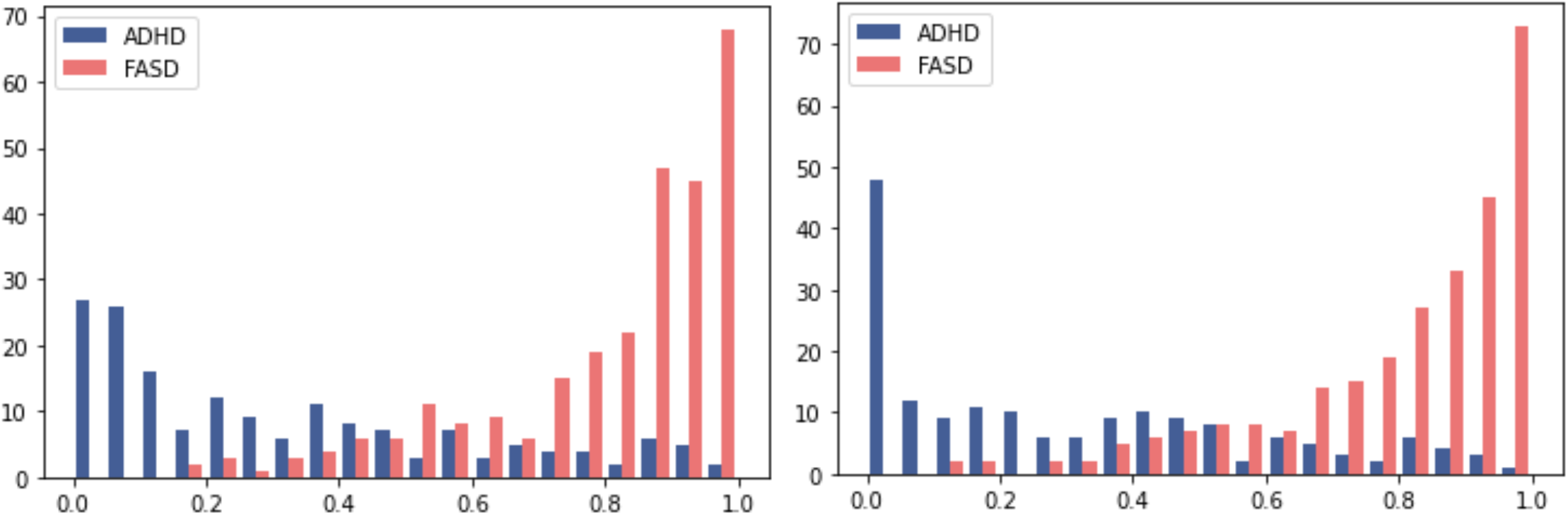
Distributions of predicted probabilities for the random forest model with 13 variables (left panel) and with 6 variables (right panel). The x-axis shows the predicted probability of having FASD and the y-axis the number of actual ADHD/FASD cases for this probability.

**Figure 3:**
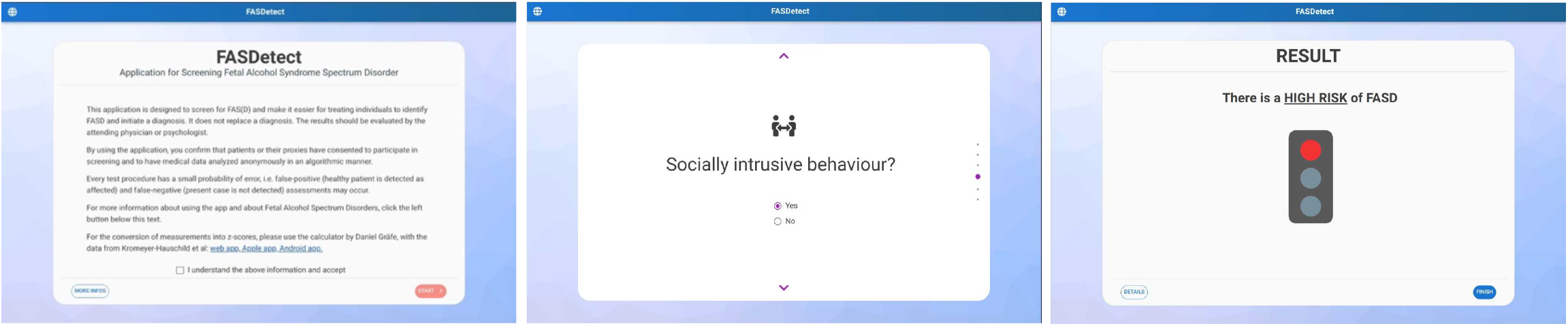
Illustration of the three screens of the FASDetect app. The first screen (left) onboards the user, the second screen (middle) contains a questionnaire asking for input to the 6 selected variables in the middle of which we show here one question on socially intrusive behaviour, and the third screen (right) shows the screening result displayed in form of a traffic light.

The training and testing of the different models contained the following steps which are described in more detail below: robust scaling, imputation, feature selection, and model fitting, all embedded in the 10-fold CV. To ensure that the contribution of each variable was similar in the prediction models, we transformed all 13 variables using robust scaling. In robust scaling, the median is subtracted from the value of each variable and each value is then divided by the interquartile range. As a second data processing step, missing values were imputed using k-nearest neighbours (kNN) imputation: each missing value was imputed using the (uniform or distance-weighted) mean value from k_i nearest neighbours found in the training set with non-missing values for the variable, where k_i is a hyperparameter of the pipeline. The distance between two points was measured by Euclidean distance, ignoring variables that were missing for either point. In the next step, we performed a variable selection among the 13 selected variables based on their estimated mutual information with the target variable. Mutual information measures the dependency between two random variables based on entropy and allows to capture also non-linear relationships. Each variable is ranked based on its mutual information with the target variable, and the highest-ranking k_f variables are selected, where k_f is a hyperparameter optimized in the pipeline. Finally, based on these transformed and quality-controlled variables, we trained and evaluated the different machine learning algorithms. In particular, we tested a logistic regression (LR), support vector machine (SVM), random forest (RF), gradient boosting decision tree (GBDT), kNN classification and Gaussian process classification algorithms. We used the lightgbm package for gradient boosting decision trees, for all other algorithms, we used the Scikit-learn implementation. Optimized hyperparameters included the number of neighbours used for imputation (k_i), the number of variables to select (k_f) or the decision whether to average values of the neighbours distance-based or uniformly for imputation. Model-specific hyperparameters for the GBDT model included the learning rate, boosting type and number of trees. For random forests, optimized model-specific hyperparameters were the minimum number of samples required to split an internal node, and the number of trees in the ensemble. For logistic regression, the regularization parameter was optimized. For support vector machines and Gaussian process classifier, the regularization parameter and kernel type were optimized. Hyperparameters optimized for the kNN classifier were the distance metric, the decision whether to average the values of the neighbours either distance-based or uniformly and the number of neighbours. The main outcome measure for the classification quality of each algorithm was the area under the receiver operating characteristic (ROC) curve (AUC), which was averaged across the 10 test datasets. The reported confidence intervals for ROC AUC scores are the average interval boundaries of confidence intervals calculated for each CV fold according to DeLong (16). In addition, we assessed the accuracy, precision, recall, and the calibration measured through the Brier score of each model. Lower Brier scores indicate better calibration (17).

In a follow-up analysis, our aim was to evaluate the performance of a most parsimonious prediction model using fewer variables, which is easier to apply in practice. To this aim, the pipeline was run again with a modified variable selection step, where only variables were selected that had been selected by at least half of the different machine learning models in at least 9 of the 10 CV trials. As described above, a variable was selected in a CV trial of an experiment with a classifier when the estimated mutual information with the target was among the k_f highest ranking features on the training set and the classifier with the best hyperparameters (including the number of variables, validated on the validation sets of this CV trial) used this variable. Six variables satisfied these criteria and were used to train the machine learning pipelines a second time.

### Ethical statement

This study was conducted in accordance with the principles of the Declaration of Helsinki and Good Clinical Practice and approved by the local ethics committee (EA2/053/20). Consent/assent requirements were waived for this retrospective chart review study.

## Data Availability

The data analyzed in this study represents sensitive identifying clinical data of patients with ADHD and patients visiting the FASD specialist center at the Campus Charite Virchow of the Charite -- Universitaetsmedizin, and cannot be made available.

## Acknowledgements

This work uses patient data collected by the staff at Charité Campus Virchow Klinikum. We would like to thank all patients and staff, who contributed and made this research possible. We are grateful to Jonathan A. Edelman and Babajide Owoyele for their help in the design of FASDetect as well as critical comments through the development.

## Author contributions

Conceptualization and methodology, OG, SK and HW; statistical analysis and implementation, LE and AW; data generation, AW, HW, OG; writing of first draft, LE and AW; supervision, SK, OG, HW, CUC; critical revision of the manuscript, all authors. All authors read and agreed on the final version of the manuscript.

## Data and code availability

The data analyzed in this study represents sensitive identifying clinical data of patients with ADHD and patients visiting the FASD specialist center at the Campus Charité Virchow of the Charité – Universitätsmedizin, and cannot be made available. All Python code used to perform the analyses is publicly available at https://github.com/HIAlab/FASDetect.

## Supplementary Figures

**Figure S1.**
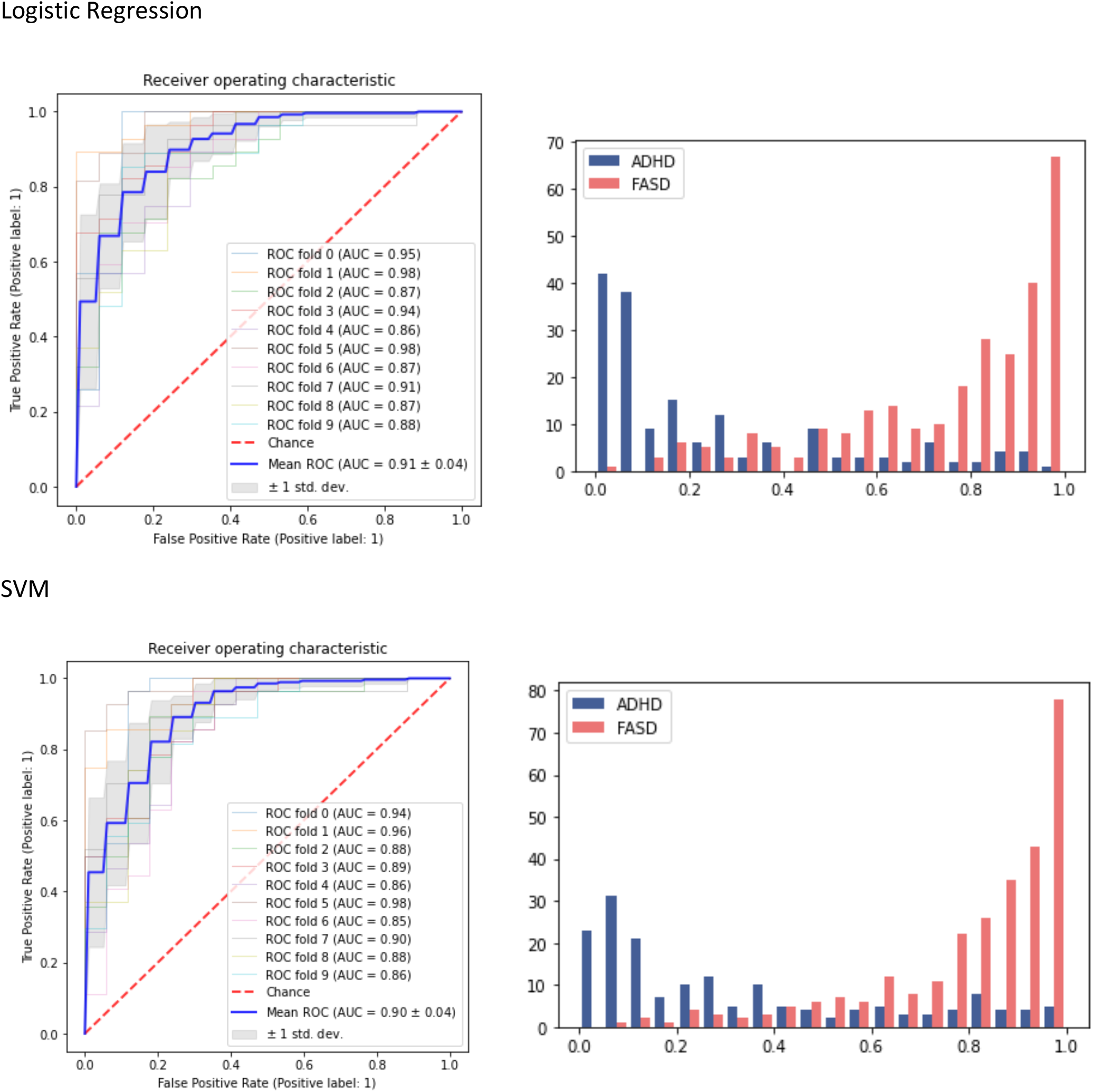
ROC curves (left column) and plot of the distribution of the predicted probabilities (right column) for logistic regression (first row) and SVM (second row), based on their application with 13 variables. In the plot of predicted probabilities, the x-axis shows the predicted probability of having FASD and the y-axis the number of actual ADHD/FASD cases for this probability.

**Figure S2.**
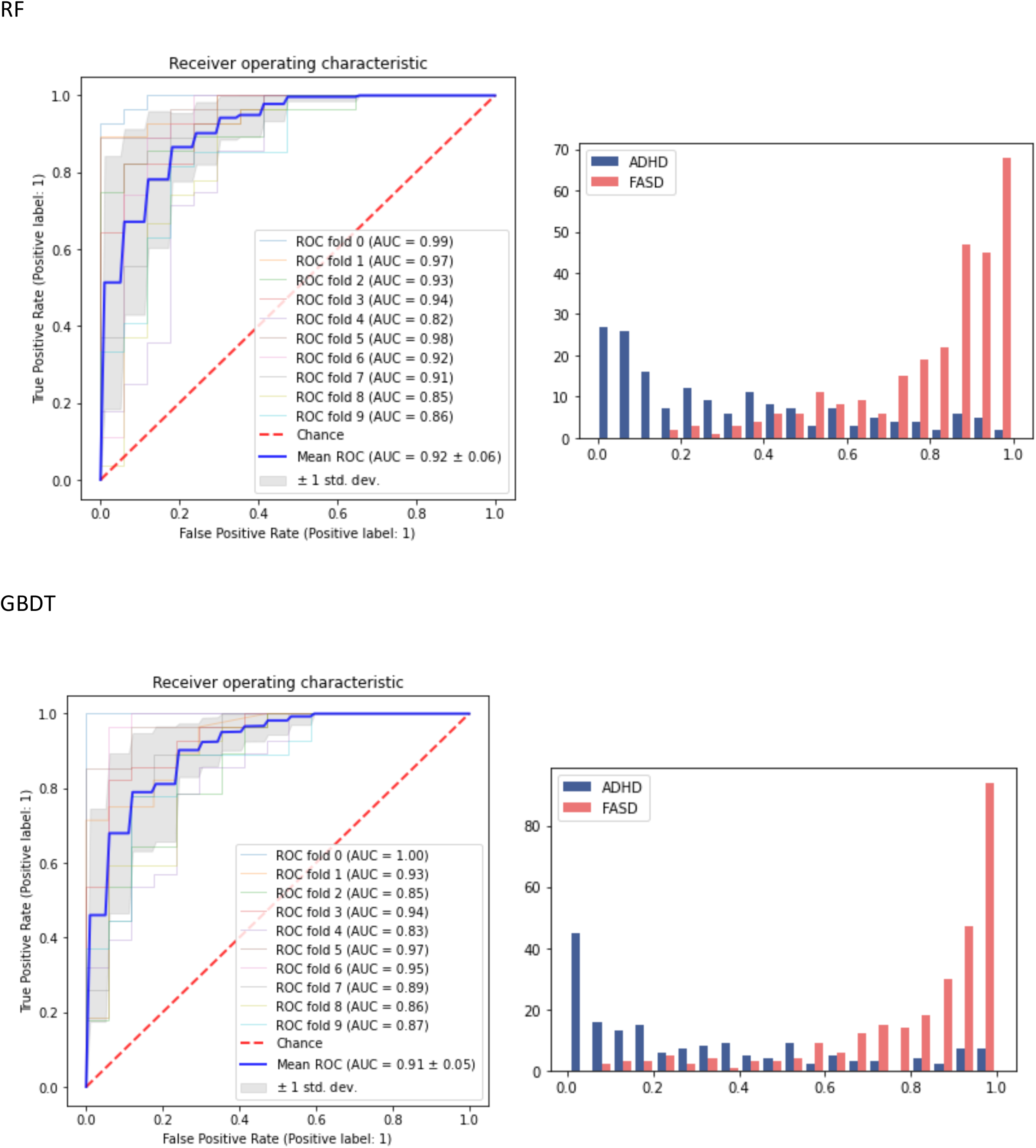
ROC curves (left column) and plot of the distribution of the predicted probabilities (right column) for random forests (first row) and GBDT (second row), based on their application with 13 variables. In the plot of predicted probabilities, the x-axis shows the predicted probability of having FASD and the y-axis the number of actual ADHD/FASD cases for this probability.

**Figure S3.**
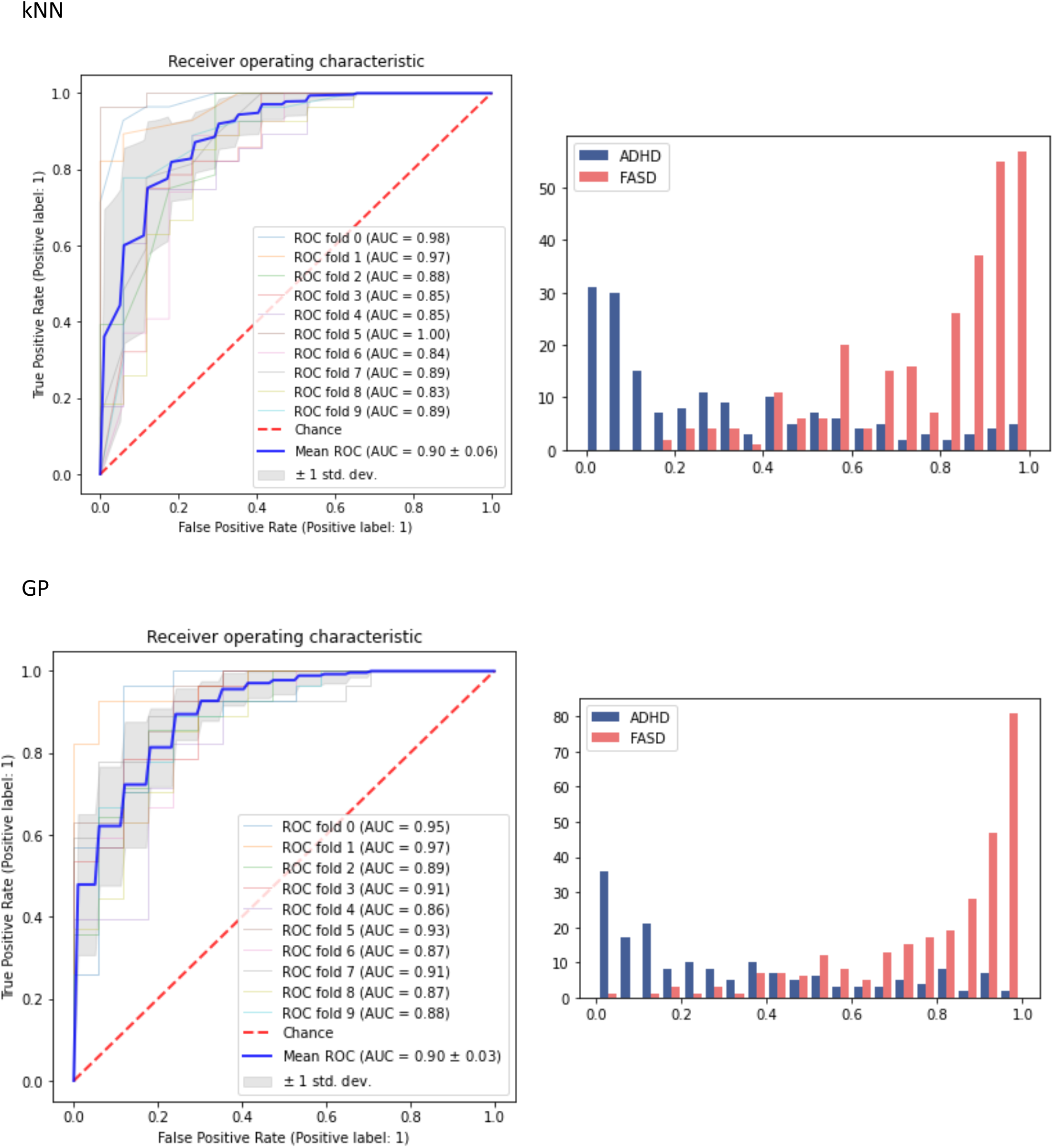
ROC curves (left column) and plot of the distribution of the predicted probabilities (right column) for kNN (first row) and GP (second row), based on their application with 13 variables. In the plot of predicted probabilities, the x-axis shows the predicted probability of having FASD and the y-axis the number of actual ADHD/FASD cases for this probability.

**Figure S4.**
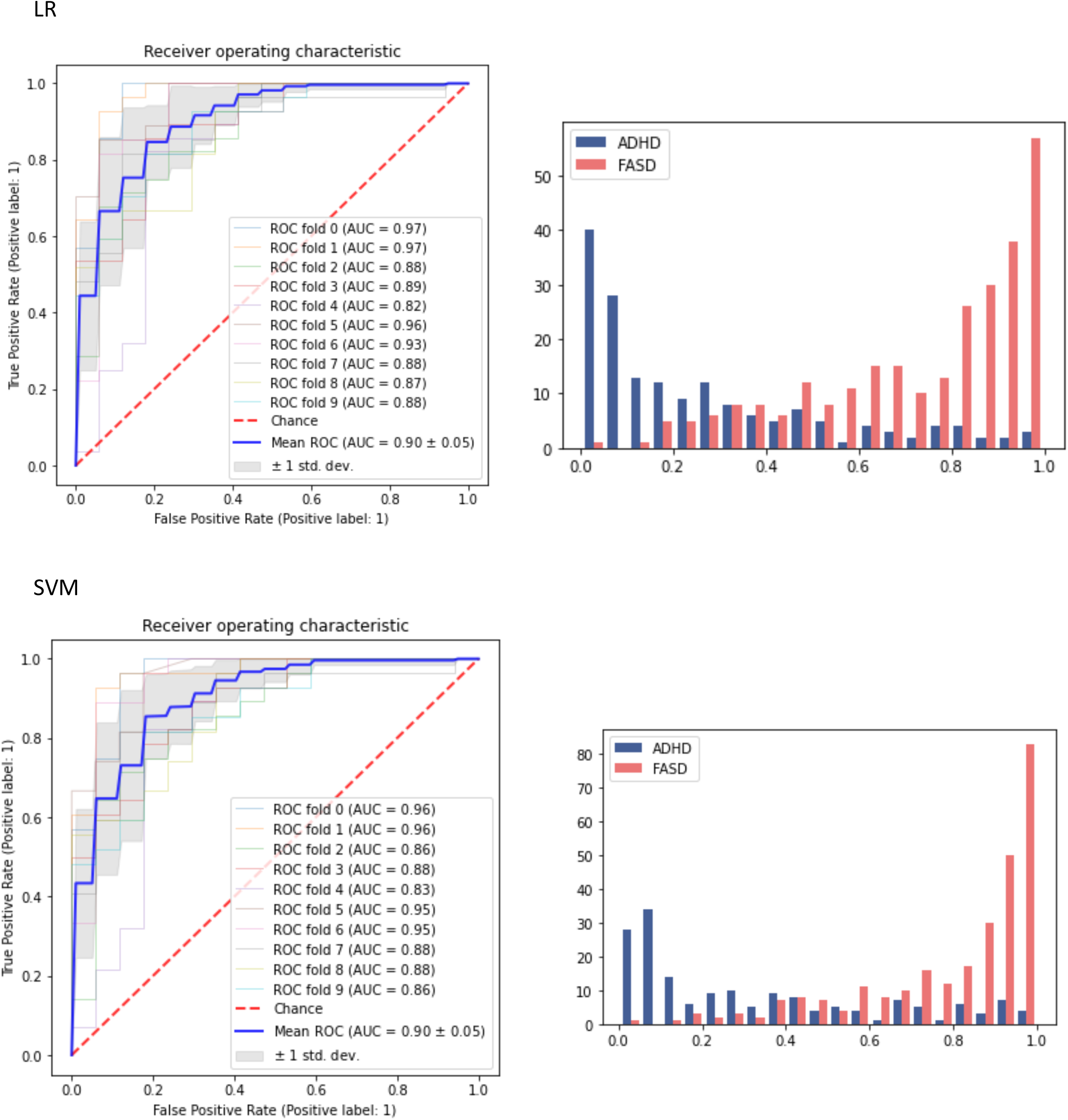
ROC curves (left column) and plot of the distribution of the predicted probabilities (right column) for logistic regression (first row) and SVM (second row), based on their application with 6 variables. In the plot of predicted probabilities, the x-axis shows the predicted probability of having FASD and the y-axis the number of actual ADHD/FASD cases for this probability.

**Figure S5.**
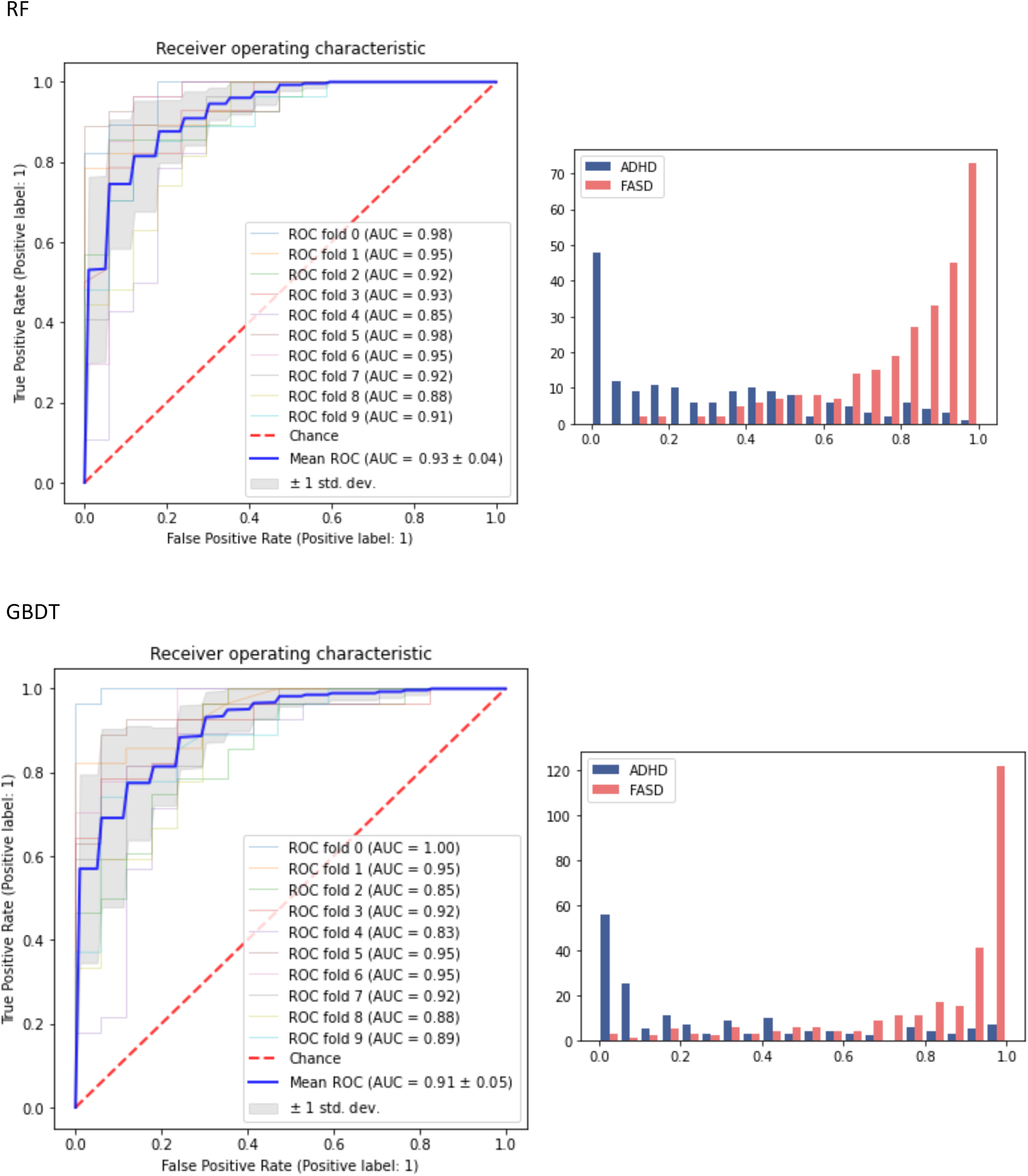
ROC curves (left column) and plot of the distribution of the predicted probabilities (right column) for random forests (first row) and GBDT (second row), based on their application with 6 variables. In the plot of predicted probabilities, the x-axis shows the predicted probability of having FASD and the y-axis the number of actual ADHD/FASD cases for this probability.

**Figure S6.**
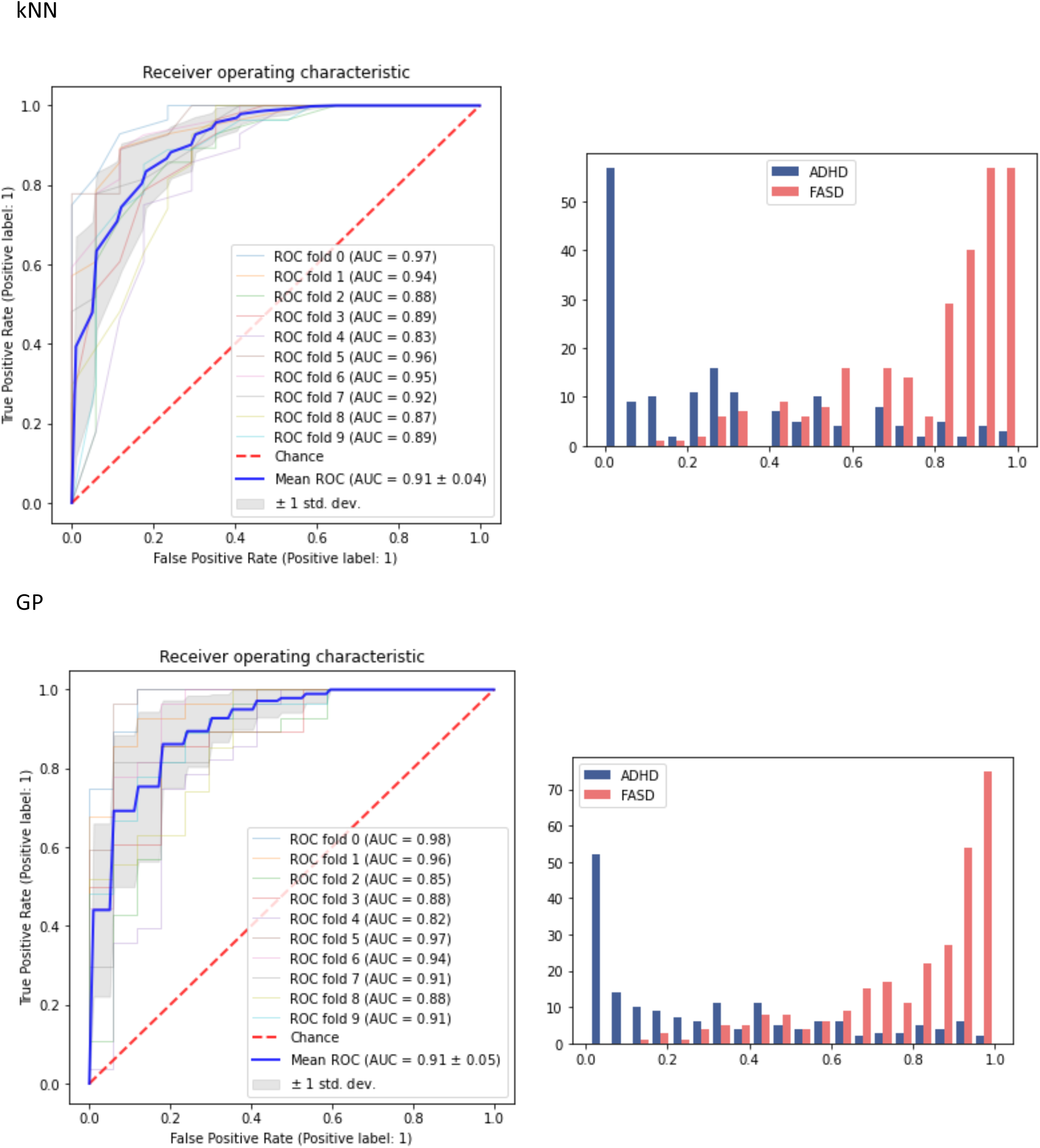
ROC curves (left column) and plot of the distribution of the predicted probabilities (right column) for kNN (first row) and GP (second row), based on their application with 5 variables. In the plot of predicted probabilities, the x-axis shows the predicted probability of having FASD and the y-axis the number of actual ADHD/FASD cases for this probability.

